# Dose–response relationship between Frailty Index and Stroke among middle-aged and elderly Chinese

**DOI:** 10.1101/2023.06.28.23292016

**Authors:** Yifang Yang, Yuxia Ma, Tingting Yang, Xiang He, Junbo Chen, Tingting Wu, Jinhan Nan, Juanjuan Feng, Lin Han

## Abstract

**Background:** Stroke has emerged as the leading cause of mortality in China, with the burden of the disease increasing with the aging population. Recent research has established a correlation between frailty and stroke, with the frailty index serving as a comprehensive measure of frailty in elderly populations. This study aimed to explore the dose-response relationship between frailty index and stroke, with the objective of providing a reference for effective stroke identification in middle-aged and elderly individuals, thereby preventing the onset of the disease.

**Methods:** The data used in this study were derived from the China Health and Retirement Longitudinal Study (CHARLS) database from 2011 to 2018. Proportional hazards model was utilized to investigate the impact of frailty index on stroke risk, while restricted cubic spline analysis was employed to examine the dose-response relationship between frailty index and stroke. Subgroup analysis was used to further understand whether the frailty index has an effect on the occurrence of stroke in different subgroups.

**Results:** A total of 11328 participants were included in the study, with 401 (3.3%) stroke patients. Frailty and pre-frailty were associated with a high risk of stroke events compared with robust group *(HR, 3*.*89, 95%CI, 2*.*88-5*.*26*), (*HR, 2*.*09, 95%CI, 1*.*60-2*.*75*), after adjusting for all covariates. There was a non-linear relationship between frailty index and stroke in the restricted cubic spline regression model. Frailty index was a risk factor of stroke when it exceeded 0.13. Taking the robust as reference, subgroup analysis results showed a significant interaction with the frailty index in all subgroup analyses except for the smoking group.

**Conclusion:** Pre-frailty and frailty were significantly associated with stroke risk. Frailty index had a nonlinear relationship with stroke occurrence, and frailty index >0.13 was significantly correlated with stroke occurrence. Frailty index is an independent predictor of stroke occurrence.

## 1. Introduction

Stroke is a cerebrovascular disease caused by the rupture or cerebral vascular obstruction, resulting in neurological dysfunction[1]. It is characterized by high incidence, high disability rate, high mortality rate, high recurrence rate, and high economic burden[2]. According to the Global Burden of Disease (GBD) study, stroke is the second leading cause of death in the world, accounting for 11.6% of total deaths[3]. In China, stroke has surpassed cardiovascular disease as the leading cause of death among residents, and the standardized incidence rate is increasing by 8.3% annually[4]. As of 2018, the per capita hospitalization costs of ischemic and hemorrhagic stroke patients in China have increased by 56% and 125%, respectively, compared to 2008. With the acceleration of the aging process, the burden of stroke is increasing[2]. Therefore, exploring the influencing factors of stroke has important clinical significance for targeted prevention.

Frailty is a multifactorial clinical syndrome associated with age, characterized by a decline in physiological reserves, disruption of self-balance, increased frailty, and reduced stress resistance[5]. It may lead to adverse health outcomes such as falls, disability, and increased risk of death[6]. With the development of global aging, frailty has gradually become an important research topic in geriatric medicine. As a comprehensive index that can effectively measure individual frailty, the frailty index is also widely used to study the health and frailty change rate of the elderly[7]. In recent years, an increasing number of studies have found a certain relationship between frailty and stroke. Some studies have found that the degree of frailty before stroke is significantly correlated with the National Institutes of Stroke Scale (NIHSS) score[6]. In Martin’s study[8], more than half of the surveyed stroke patients were considered to be in the pre-frailty. However, most studies currently focus on exploring the impact of frailty on stroke outcomes[9, 10], limited studies have assessed the relationship between the frailty index and the risk of stroke events, the correlation between the frailty index of middle-aged and elderly people in China and stroke has not been reported. But exploring the dose-response relationship can help quantify the correlation between frailty index and stroke risk, and provide a better reference for stroke prevention. Therefore, the aim of this study was to investigate the relationship between the frailty index and stroke, and to provide a reference for effective frailty screening and stroke identification in middle-aged and elderly people to prevent the occurrence of the disease.

## 2 Materials and Methods

### 2.1 Participants

The data used in this study were derived from the China Health and Retirement Longitudinal Study (CHARLS) database from 2011 to 2018[10]. The survey targeted Chinese middle-aged and elderly people aged 45 and above. The survey covered 28 provinces (autonomous regions, municipalities), 150 counties, and 450 communities (villages) in China, and collected information on sociodemographic characteristics and personal health status. CHARLS is a nationally representative dataset with rich information. The study was approved by the Institutional Review Board of Peking University (IRB00001052-11015), and written informed consent was obtained from all participants or their legal representatives.

In this study, data from CHARLS in 2011, 2013, 2015, and 2018 were utilized. Participants who met the following criteria were excluded: (1) age < 45 years; (2) diagnosed with stroke at baseline; (3) incomplete survey data. Finally, a total of 11328 participants aged over 45 years were included in the longitudinal analysis. The selection process of individuals included in the study is shown in Figure 1.

**Figure 1.**
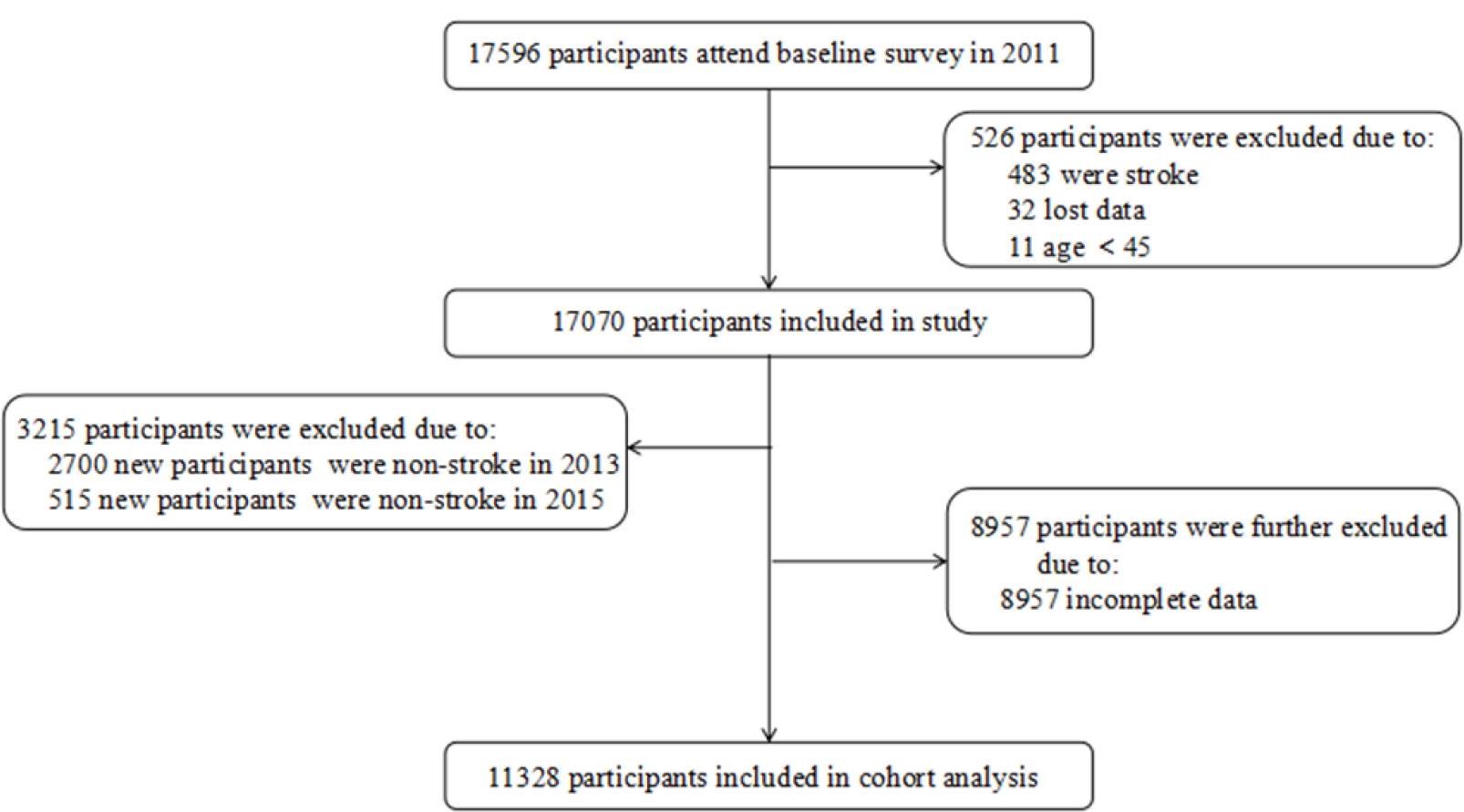
Flowchart of study participants.

### 2.2 Measurement

#### 2.2.1 Frailty Index

The cumulative deficit model developed by Rockwood et al. was employed in this study and created according to the standard process of frailty index[11]. The variables included in the model needed to satisfy the following three conditions: (1) the deficit must be age-related; (2) the prevalence of the deficit must increase with age;(3) the deficit cannot saturate too early. Based on previous studies [12, 13]and the CHARLS questionnaire catalog, a total of 36 deficits were included, encompassing self-assessment, activities of daily living, chronic diseases, and mental health. Each item was scored between 0 and 1, with 0 indicating the absence of a deficit and 1 indicating the presence of a deficit. Given that some items have more than two answer options (e.g., how would you rate your health? very good, good, fair, poor, very poor), the scores were recoded (very good = 0, good = 0.25, fair = 0.5, poor = 0.75, very poor = 1). The frailty index was calculated by summing the number of deficits present in the study subjects and dividing by the total number of deficits[14].Based on previous research, three states were defined: robust (frailty index ≤ 0.10), prefrail (frailty index > 0.10 to < 0.25), and frail (frailty index ≥ 0.25).

#### 2.2.2 Outcomes

The primary outcome measure for this study was stroke. The outcome was mainly given by the questionnaire “Have you been diagnosed with Stroke by a doctor?” “When did you first diagnose or know of this situation ?” “Since the last visit / in the last two years, has your doctor told you that you had another stroke ?” to evaluate. The affirmative answer was considered a stroke.

#### 2.2.3. Covariates

Based on previous research[15, 16], the covariates included in the study are demographic and health-related factors. Demographic variables consist of age, gender, marital status (married, divorced, widowed, single), education level (primary school, middle school, high school / vocational school, college / university or above), and place of residence (urban, suburban, rural, special areas). Health-related factors comprise smoking, drinking, hypertension, diabetes, dyslipidemia, glycated hemoglobin (HbA1c), triglycerides (TG), low-density lipoprotein cholesterol (LDL-c), total cholesterol (CHO), creatinine (CREA).

### 2.3 Quality control

The response rate and data quality of the CHARLS questionnaire are among the highest in similar surveys worldwide. The questionnaire design was informed by international experience, including the United States of America Health and Retirement Study, The United Kingdom of Great Britain and Northern Ireland ‘s English Longitudinal Study of Ageing, and the European Health, Ageing and Retirement Study. The questionnaire underwent structured modification and design after pre-survey testing. During the survey implementation, a quality control team was established to ensure standardized and regulated data collection through structured survey forms, simulated training for survey teams, real-time quality checks and feedback, intervention in data collection behavior of surveyors, GPS positioning, audio recording checks, and telephone verification. These measures ensured the standardization and regulation of data collection.

### 2.4 Statistical Analysis

Continuous variables were characterized using mean (standard deviation, SD) or median (interquartile range, IQR), while categorical data were described using frequencies (%). Inter-group comparisons were conducted using ANOVA or Kruskal-Wallis for continuous variables and χ 2 test for categorical data. Proportional hazards model was utilized to investigate the impact of frailty index on stroke risk, while restricted cubic spline analysis was employed to examine the dose-response relationship between frailty index and stroke.

## 3 Results

### 3.1 Participant Characteristics

A total of 11328 participants were included in the study and detailed characteristics are shown in Table 1. The mean age of the participants was 66.79 ± 12.49, of which 5470 were males, the prevalence of pre-frailty and frailty was respectively was 42.96 % and 21.17 %. The incidence of stroke was 3.3%. The median frailty index for all participants was 0.13( 0.08, 0.21). There were significant differences in age, education level, marital status, residence, drinking, hypertension, diabetes, dyslipidemia, stroke, HbA1c, CHO, LDL-c, CREA, and TG among the three groups of robust, pre-frailty, and frailty.

**Table 1.**
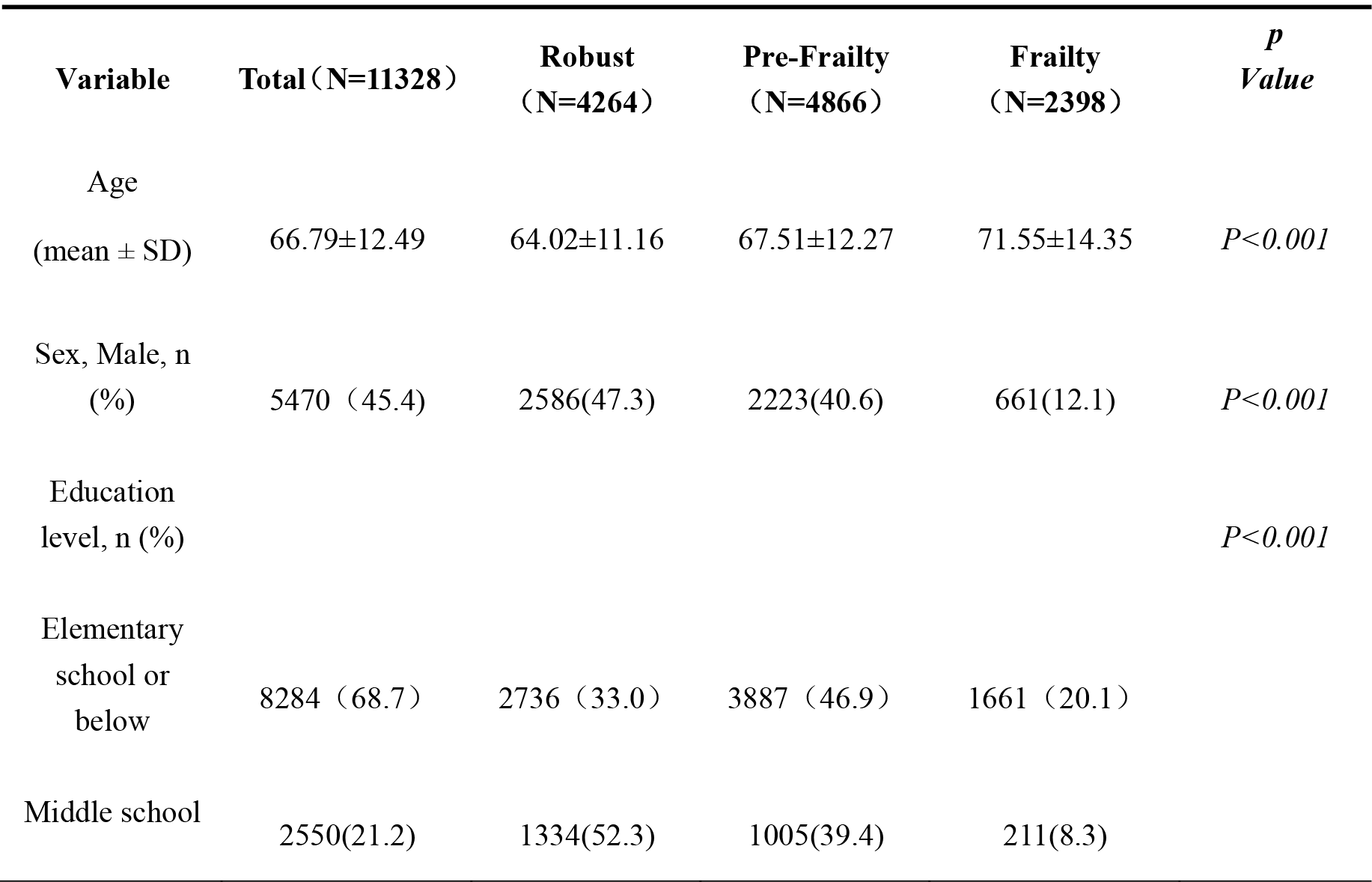
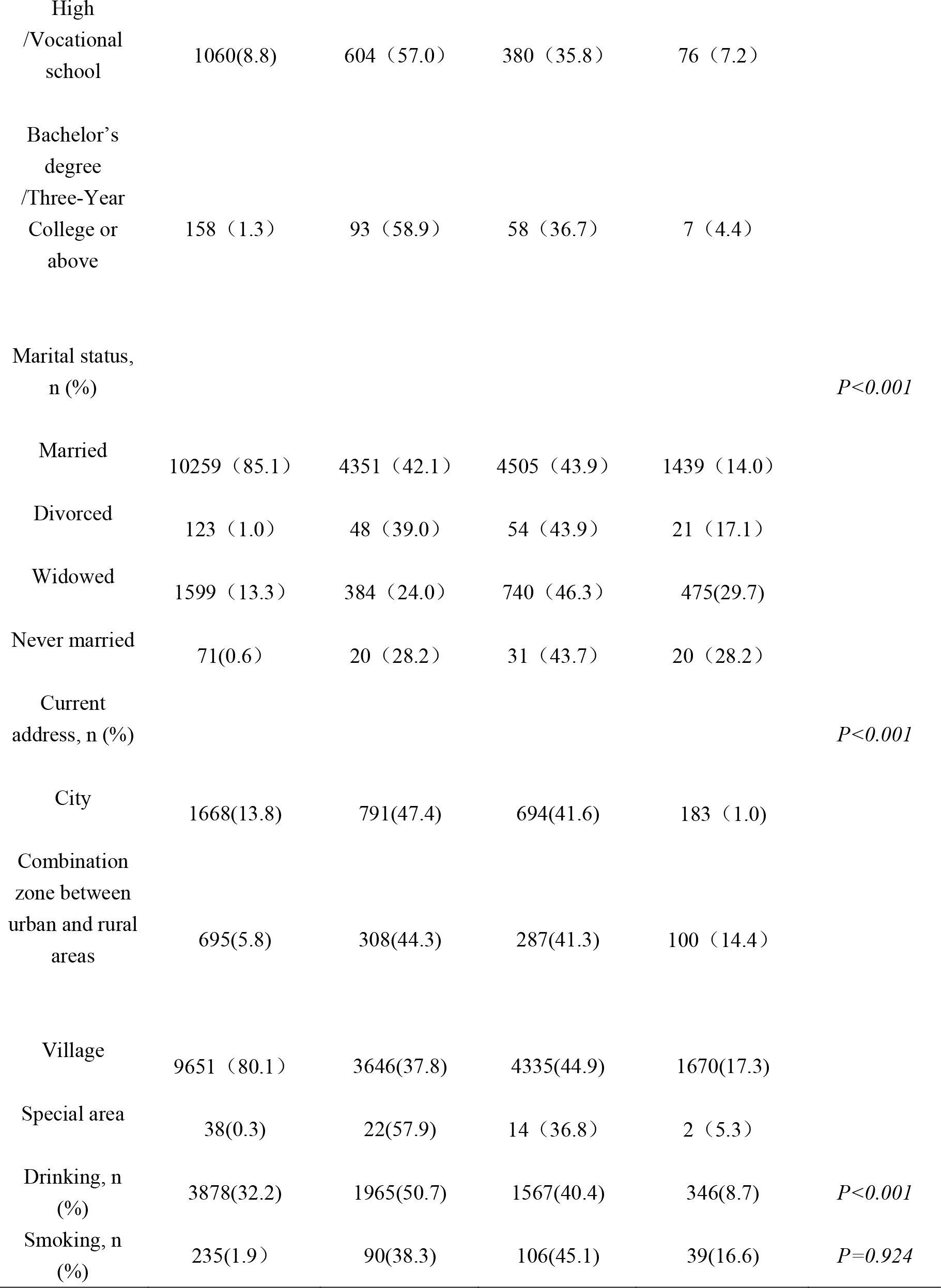
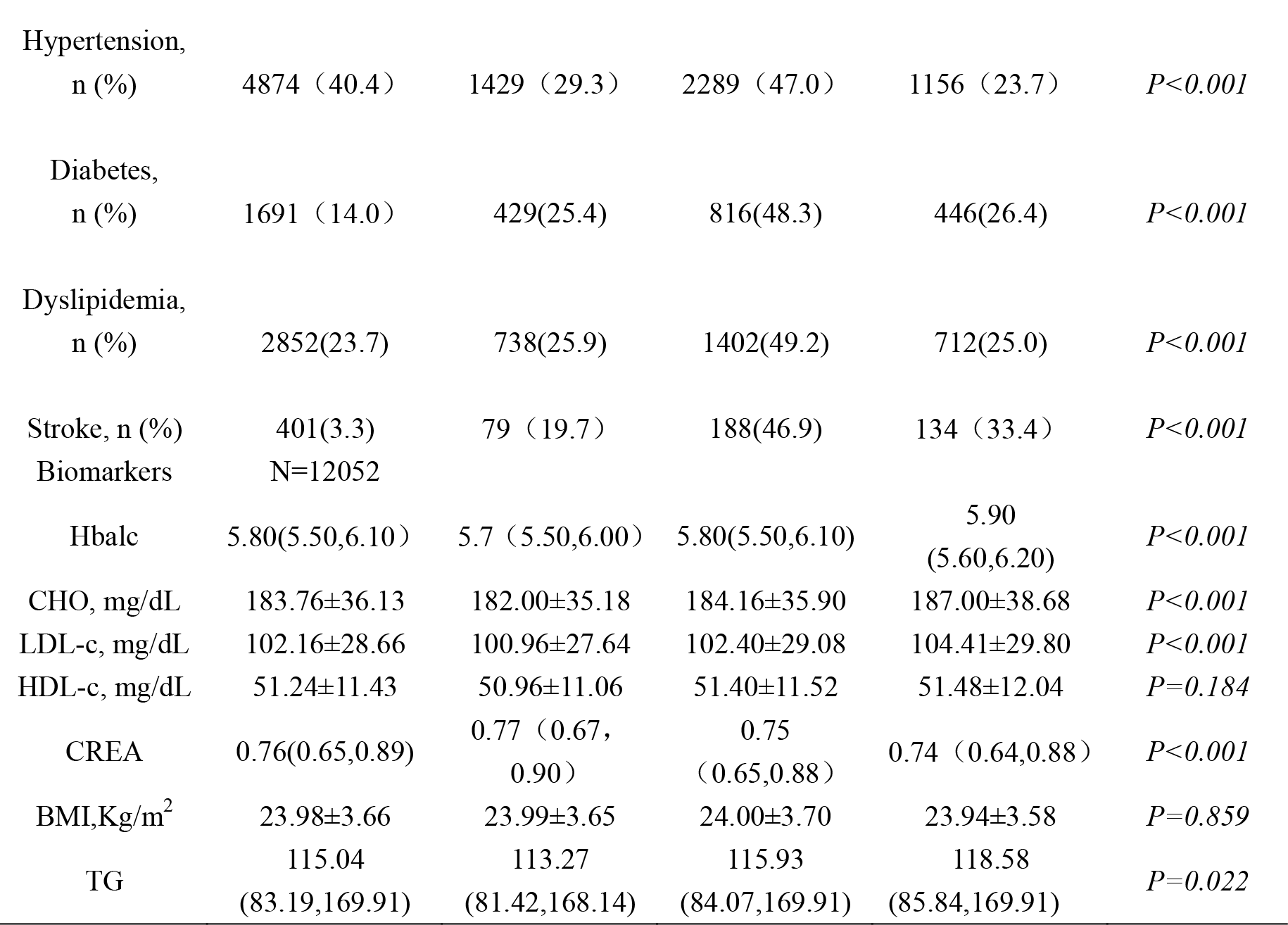
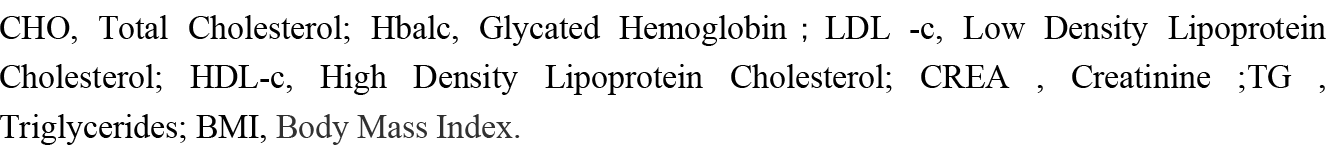
Characteristics of study population by frailty status.

| Variable | Total (N=11328) | Robust<br>(N=4264) | Pre-Frailty<br>(N=4866) | Frailty<br>(N=2398) | *P*<br>Value |
| --- | --- | --- | --- | --- | --- |
| Age<br>(mean ± SD) | 66.79±12.49 | 64.02±11.16 | 67.51±12.27 | 71.55±14.35 | $P<0.001$ |
| Sex, Male, n<br>(%) | 5470 (45.4) | 2586(47.3) | 2223(40.6) | 661(12.1) | $P<0.001$ |
| Education<br>level, n (%) | | | | | $P<0.001$ |
| Elementary<br>school or<br>below | 8284 (68.7) | 2736 (33.0) | 3887 (46.9) | 1661 (20.1) |  |
| Middle school | 2550(21.2) | 1334(52.3) | 1005(39.4) | 211(8.3) |  |

| Variable | Total (N=11328) | Robust<br>(N=4264) | Pre-Frailty<br>(N=4866) | Frailty<br>(N=2398) | *P*<br>*Value* |
| --- | --- | --- | --- | --- | --- |
| High<br>/Vocational<br>school | 1060(8.8) | 604 (57.0) | 380 (35.8) | 76 (7.2) |  |
| Bachelor's<br>degree<br>/Three-Year<br>College or<br>above | 158 (1.3) | 93 (58.9) | 58 (36.7) | 7 (4.4) |  |
| Marital status,<br>n (%) |  |  |  |  | *P*<0.001 |
| Married | 10259 (85.1) | 4351 (42.1) | 4505 (43.9) | 1439 (14.0) |  |
| Divorced | 123 (1.0) | 48 (39.0) | 54 (43.9) | 21 (17.1) |  |
| Widowed | 1599 (13.3) | 384 (24.0) | 740 (46.3) | 475(29.7) |  |
| Never married | 71(0.6) | 20 (28.2) | 31 (43.7) | 20 (28.2) |  |
| Current<br>address, n (%) |  |  |  |  | *P*<0.001 |
| City | 1668(13.8) | 791(47.4) | 694(41.6) | 183 (1.0) |  |
| Combination<br>zone between<br>urban and rural<br>areas | 695(5.8) | 308(44.3) | 287(41.3) | 100 (14.4) |  |
| Village | 9651 (80.1) | 3646(37.8) | 4335(44.9) | 1670(17.3) |  |
| Special area | 38(0.3) | 22(57.9) | 14 (36.8) | 2 (5.3) |  |
| Drinking, n<br>(%) | 3878(32.2) | 1965(50.7) | 1567(40.4) | 346(8.7) | *P*<0.001 |
| Smoking, n<br>(%) | 235(1.9) | 90(38.3) | 106(45.1) | 39(16.6) | *P*=0.924 |
| Hypertension,<br>n (%) | 4874 (40.4) | 1429 (29.3) | 2289 (47.0) | 1156 (23.7) | *P*<0.001 |
| Diabetes,<br>n (%) | 1691 (14.0) | 429(25.4) | 816(48.3) | 446(26.4) | *P*<0.001 |
| Dyslipidemia,<br>n (%) | 2852(23.7) | 738(25.9) | 1402(49.2) | 712(25.0) | *P*<0.001 |
| Stroke, n (%) | 401(3.3) | 79 (19.7) | 188(46.9) | 134 (33.4) | *P*<0.001 |
| Biomarkers | N=12052 |  |  |  |  |
| HbA1c | 5.80(5.50,6.10) | 5.7 (5.50,6.00) | 5.80(5.50,6.10) | 5.90<br>(5.60,6.20) | *P*<0.001 |
| CHO, mg/dL | 183.76±36.13 | 182.00±35.18 | 184.16±35.90 | 187.00±38.68 | *P*<0.001 |
| LDL-c, mg/dL | 102.16±28.66 | 100.96±27.64 | 102.40±29.08 | 104.41±29.80 | *P*<0.001 |
| HDL-c, mg/dL | 51.24±11.43 | 50.96±11.06 | 51.40±11.52 | 51.48±12.04 | *P*=0.184 |
| CREA | 0.76(0.65,0.89) | 0.77 (0.67,<br>0.90) | 0.75<br>(0.65,0.88) | 0.74 (0.64,0.88) | *P*<0.001 |
| BMI,Kg/m<sup>2</sup> | 23.98±3.66 | 23.99±3.65 | 24.00±3.70 | 23.94±3.58 | *P*=0.859 |
| TG | 115.04<br>(83.19,169.91) | 113.27<br>(81.42,168.14) | 115.93<br>(84.07,169.91) | 118.58<br>(85.84,169.91) | *P*=0.022 |

CHO, Total Cholesterol; HbA1c, Glycated Hemoglobin ; LDL -c, Low Density Lipoprotein Cholesterol; HDL-c, High Density Lipoprotein Cholesterol; CREA , Creatinine ;TG , Triglycerides; BMI, Body Mass Index.

CHO, Total Cholesterol; Hbalc, Glycated Hemoglobin;LDL -c, Low Density Lipoprotein Cholesterol; HDL-c, High Density Lipoprotein Cholesterol; CREA, Creatinine ;TG, Triglycerides; BMI, Body Mass Index.

### 3.2 Association between the Frailty Index and Stroke

Table 2 shows the relationship between the frailty index and stroke, with all models for stroke satisfying the proportional hazards assumption. Of the 11,328 individuals included, 401 had a stroke. Frailty and pre-frailty were associated with a high risk of stroke events compared with robust group, after adjusting for age, sex, education level, location and marital status, smoking, drinking, hypertension, diabetes, dyslipidemia, HbAlc, cho,LDL-C,HDL-C, CREA, BMI, tg, with HRs of *3*.*89 (2*.*88,5*.*26), 2*.*09 (1*.*60,2*.*75)*. Each 0.1 increment in the frailty index was associated with a 27% increase in the risk of stroke *(HR,1*.*28,95%CI,1*.*12-1*.*47*) (Table 2).

**Table 2.** Longitudinal association between the frailty index and Stroke.

| Outcome | Case/N | Model 1 | Model 2 | Model 3 |
| --- | --- | --- | --- | --- |
|  |  | HR(95%CI) | HR(95%CI) | HR(95%CI) |
| Stroke |  |  |  |  |
| Robust | 4264/11328 | Reference | Reference | Reference |
| Pre-frailty | 4866/11328 | 2.41(1.85,3.14) | 2.09(1.60,2.73) | 2.09(1.60,2.75) |
| Frailty | 2398/11328 | 5.41(4.08,7.19) | 3.82(2.83,5.16) | 3.89(2.88,5.26) |
| Per 0.1 increment |  | 1.43(1.34,1.53) | 1.28(1.12,1.47) | 1.28(1.12,1.47) |

Model 1 was adjusted age and gender. Model 2 was additionally adjusted educational level, marital status, residence, smoking status, drinking status, hypertension, diabetes, dyslipidemia. Model 3 was additionally adjusted HbA1c, TG, LDL-C, CHO, CREA.

### 3.3 The dose-response relationship between frailty index and stroke

Figure 2 shows the dose-response relationship between frailty index and stroke. The restricted cubic spline shows a non-linear relationship between frailty index and stroke, frailty index > 0.13 was associated with the high risk of stroke events and we found that the risk of stroke increased with increasing frailty index.

**Figure 2.**
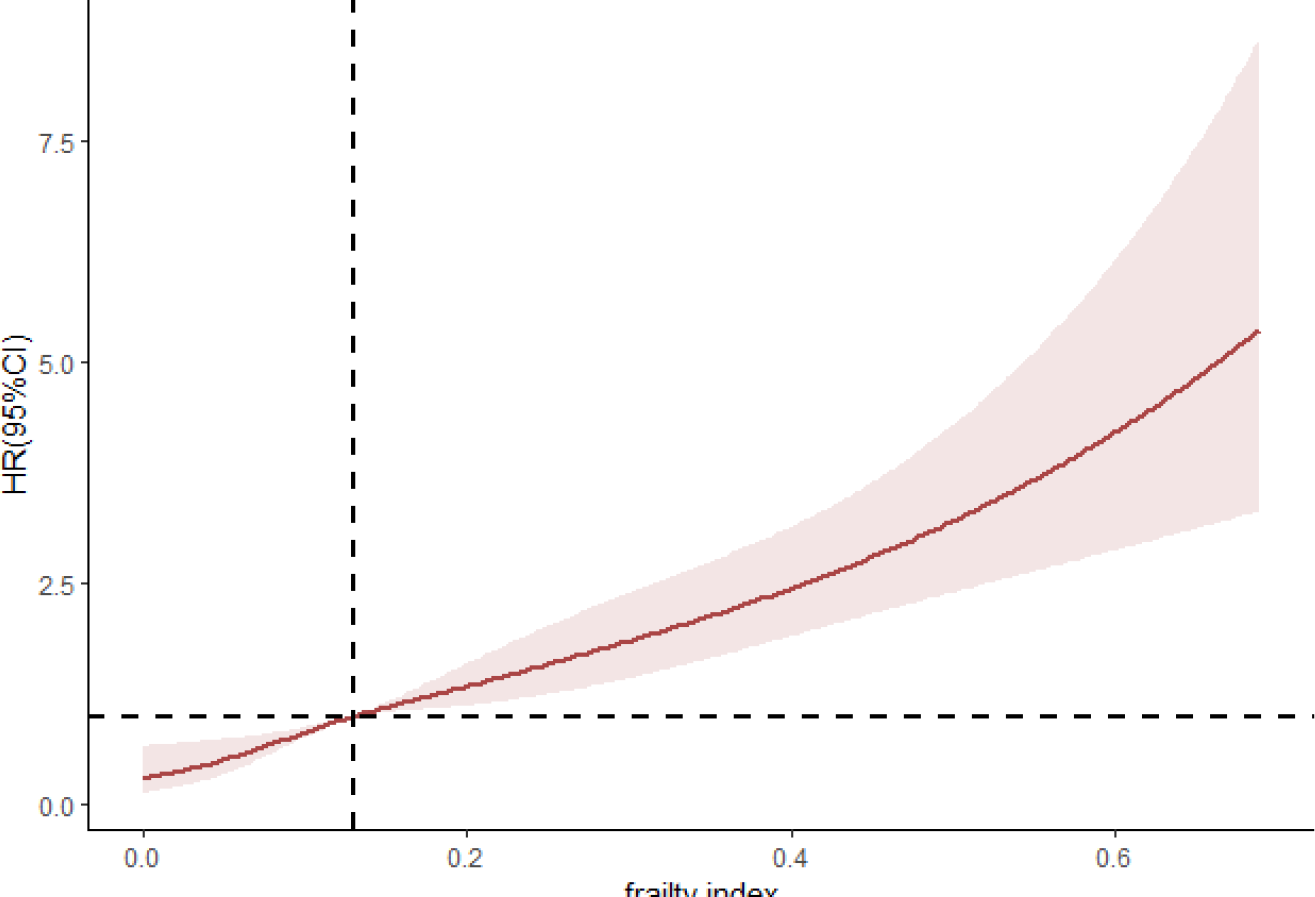
The dose-response relationship between frailty index and stroke.

### 3.4 Subgroup Analysis

To further understand whether the frailty index has an effect on the occurrence of stroke in different subgroups, we performed a subgroup analysis based on the demographic characteristics of the participants. Taking the robust as reference, the results showed a significant interaction with the debilitating index in all subgroup analyses except for the smoking group (Figure 3).

**Figure 3.**
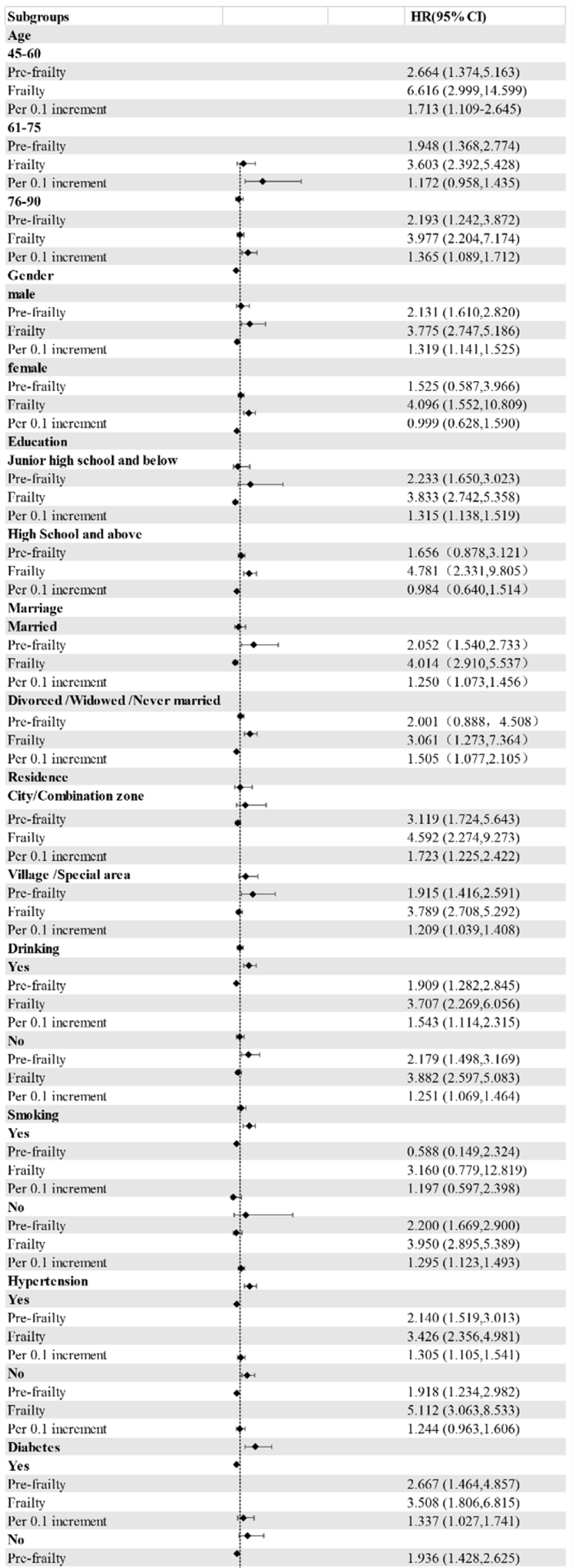
Forest plot of frailty index of the with the risk of stroke.

## 4 Discussion

This study represents the first cohort analysis of a large, nationally representative cohort to investigate the association between frailty index and stroke risk in middle-aged and older adults in China. Frailty index was utilized to define frailty, and the results indicated that 33.4% of stroke patients were frailty. This finding contrasts with the results of Taylor-Rowan et al[8], who conducted a cross-sectional study in a hospital setting to examine the prevalence of pre-stroke frailty in stroke patients, which was 28%. This difference may be attributed to the larger sample size and the inclusion of more patients with poorer predisease function in this study. Furthermore, the two studies employed different methodologies. These different study designs may have contributed to the observed differences in results.

Furthermore, this study highlights the importance of frailty assessment in midlife as a means of preventing the development of late stroke. The average age of the frailty group was higher than that of the robust group and the pre-frailty period, and frailty increased with age, consistent with previous research[17]. However, the subgroup analysis revealed that the frailty index was the strongest predictor of stroke occurrence in the population aged 45-65 years. This finding is in line with previous studies[14, 18], indicating that assessing frailty during midlife can aid in preventing the onset of stroke later in life.

Our findings suggest that the frailty index may not be a reliable predictor of stroke occurrence in women, which contrasts with previous research[18], Collard et al. [19]reported a significantly higher prevalence of frailty in women (9.6%, 95% *CI* = *9*.*2-10*.*0%*) than in men (5.2%, 95% *CI = 4*.*9-5*.*5%. X*^*2*^ = *298*.*9* df = 1, *P 0<*.*001*). Given the bidirectional relationship between frailty and stroke[20], frailty is considered an influencing factor of stroke, and gender makes a difference in stroke events. Therefore, there may be an interaction between gender, frailty index, and stroke. There were fewer strokes in the women in our study, so the results may have been accidental. As the population of older adults continues to grow, understanding the relationship between stroke and frailty in women becomes increasingly pressing.

Additionally, our study raises the possibility that the association between frailty intensity and stroke may be stronger in urban areas and among individuals with lower education levels. However, given the limited research on the relationship between frailty index, residence, education level, and stroke, it is necessary that further studies be conducted to elucidate the potential mechanisms underlying these associations.

Currently, few studies have explored the relationship between frailty index and stroke, although previous studies have identified frailty as a risk factor for stroke[21]. In our study, we utilized frailty index as a continuous variable and analyzed 7-year follow-up data from CHARLS. After fully adjusting for traditional stroke risk factors, we observed a non-linear relationship between frailty index and stroke, with a frailty index > 0.13 being associated with a high risk of stroke events. Our findings suggest that frailty index is an independent predictor of stroke events.

The findings of this study hold significant implications for the screening of frailty and identification of disease in middle-aged and elderly populations. The results provide valuable insights for health management and intervention measures aimed at mitigating the risk of stroke in at-risk individuals, thereby preventing the onset of disease.

However, this study has some limitations. Firstly, the diagnosis of stroke was based on participants’ self-reported recall of a doctor’s diagnosis, which may be subject to recall bias. Secondly, while we controlled for traditional stroke risk factors, there may be other risk factors that were not accounted for. Thirdly, the inclusion of frailty index was determined by CHARLS, and the inclusion of mental health was limited, which may have introduced some degree of bias in our findings. Therefore, it is suggested that future studies should improve the existing limitations and further explore the influence of changes in the trajectory of frailty index on stroke.

## 5 Conclusions

In conclusion, we found that pre-frailty and frailty were significantly associated with stroke risk over a 7-year follow-up period in this study, even after controlling for traditional stroke risk factors. A non-linear relationship between frailty index and stroke occurrence was found by restricted cubic spline plots, with a frailty index >0.13 being significantly associated with stroke occurrence. The frailty index is an independent predictor of stroke occurrence. Therefore, with the progression of population aging, frailty assessment is recommended as part of a stroke occurrence risk prevention program for older adults to identify and prevent stroke occurrence and progression.

## Data Availability

The data used in this study were derived from the China Health and Retirement Longitudinal Study (CHARLS) database from 2011 to 2018, The study was approved by the Institutional Review Board of Peking University (IRB00001052-11015), and written informed consent was obtained from all participants or their legal representatives.

